# Prevalence of low back pain among working Ethiopian population: A systematic review and meta-analysis

**DOI:** 10.1101/2020.11.29.20238170

**Authors:** Amanuel Godana Arero, Godana Arero, Shimels Hussien Mohammed, Sahar Eftekhari

**Affiliations:** Students’ Scientific Research Center, Tehran University of Medical Sciences, Tehran, Iran; Department of Public Health, Adama Hospital Medical College, Adama, Ethiopia; Department of Community Nutrition, School of Nutritional Sciences and Dietetics, Tehran University of Medical Sciences, Tehran, Iran; School of Medicine- International Campus, Tehran University of Medical Sciences (TUMS), Tehran, Iran

**Keywords:** Low back pain, Epidemiology, Occupational injury, Working population, Ethiopia

## Abstract

**Background and objective:** Low back pain (LBP) as musculoskeletal disorder remains a common health problem and is one of the most prevalent occupational injuries affecting adults living in both developed and developing countries. To increase the power and improve the estimates of the prevalence of LBP among the working Ethiopian population, a comprehensive meta-analysis was carried out.

**Methods:** A comprehensive systematic literature search was conducted in multiple international electronic bibliographic databases such as Web of Science, Pub Med, EMBASE, Scopus, and Google Scholar. Population-based Studies into the Prevalence of LBP among the working population living in Ethiopia were included. Methodological quality for included studies was appraised using an adapted tool. Meta-analyses, Meta-regression, and sensitivity analysis were conducted. Funnel plot symmetry visualization followed by Begg’s rank correlation and Egger’s regression asymmetry test methods were performed to detect the existence of publication bias. Heterogeneity between studies was assessed by using the Cochrane Q and I2-statistics.

**Results:** In all 719 articles were identified and 13 articles with 6513 participants met the inclusion criteria for meta-analyses after filtering. The pooled point and twelve-month prevalence of LBP among working Ethiopian population was 49% (95% CI 40; 58) and 56% (95% CI 49; 62) respectively.

**Conclusion:** The results showed a high prevalence of LBP among working Ethiopian population, especially among Teachers. We believe that Prevention strategies addressing the early onset of LBP among the working population would most likely be the answer to the burden of LBP on future economies in Ethiopia.

## Introduction

It is believed that Lower back pain (LBP) is one of the most prevalent musculoskeletal conditions affecting adults living in both developed and developing nations (1-4). Broadly defined as pain or discomfort in the lumbar region of the spine between the lower costal margins and the inferior gluteal folds with or without leg pain (i.e., sciatica) (1-4). LBP remains one of the most common health problems that 50–80% of adults of working age population experience at some point in the course of their lifetime and its prevalence or incidence has been found to increase with an increase in age(3, 4). It is estimated that 46% of workers in European countries(5) and about 20% of workers in US (6, 7) report LBP at any given time. LBP is the major cause of work absence and activity restriction throughout much of the world, imposing a high economic burden on individuals, families, communities, industries and governments(8, 9).Direct costs for LBP in US are estimated between $20 billion and $98 billion, with indirect annual costs included the total cost estimates are as high as $200 billion(10, 11).

According to the Global Burden of Disease (GBD) 2010 study, the overall burden of LBP arising from workplace exposure was estimated at 21.8 million [95% CI (14.5–30.5)] disability-adjusted life years (DALYs) and is currently, the sixth-highest burden and is the cause of more years lived with disability (YLDs) globally than any other condition (12-14). Despite the fact that the literature on the prevalence of low back pain is accumulating, but for the most part studies are restricted and only available for developed countries, which comprise less than 20% of the world’s population. Understanding the prevalence of LBP in the working population in developing countries such as Ethiopia may assist in the understanding of the global LBP burden and its management. Thus, the purpose of this review aims to systematically appraise published Disease prevalence studies conducted in Ethiopia and estimate LBP prevalence in the working population, in order to ascertain whether LBP is of concern in Ethiopia, as it is globally.

## Methods

This systematic review and meta-analysis was conducted Based on the recommendations of Meta-analysis of Observational Studies in Epidemiology guideline(15), and PRISMA(Preferred Reporting Items for Systematic Reviews and Meta-Analysis) guideline (16).The work was registered in PROSPERO(protocol registration number: CRD42020188523).

### Characteristics of studies

All studies conducted in the field of the prevalence of LBP in Ethiopia, regardless of the publication period, we reviewed and included in our study. Studies could report on the prevalence of musculoskeletal disorders as a whole, yet had to provide subgroup data for the prevalence of LBP. Subjects included in the studies could be any race, gender, and age. All Studies included are published in English.

### Data sources

A systematic literature search was conducted in multiple international electronic bibliographic databases such as Web of Science, Pub Med, EMBASE, Scopus, and Google Scholar until 2020. The main keywords used for this search were: low back pain, musculoskeletal pain, occupational injury, prevalence, and Ethiopia. It should also be noted that articles published in journals and/or Dissertations/theses, conference proceedings, reports, commentaries/letters, and other grey literature and all other references to the relevant articles were included in our search. The First Reviewer (AGA) searched independently and the second reviewer (GA) checked the finding.

### Study selection

Inclusion criteria dealt with cross-sectional studies that present the prevalence of LBP among Ethiopia working population and conducted in Ethiopia. Exclusion criteria included irrelevant studies, articles without adequate data regarding observations, studies that linked LBP with other diseases, duplicate studies and Dissertations, conference proceedings, commentaries/letters, and other grey literature were excluded from this review.

### Data extraction and study quality assessment

Data extraction and quality assessment were done by two independent reviewers (AGA and GA). Cases of a discrepancy were resolved by consensus between two reviewers. For all included articles, a form was designed using a data extraction sheet in EXCEL software that include the following variables: Author names, city/region, study design, year of publication, study setting, data collection period, population, age, gender, working experience, Body mass index, type of standard questionnaire, sample size, responsive rate, the prevalence of LBP, and study quality score. The methodological quality critical appraisal tool used in a systematic and meta-analysis into the prevalence of LBP in Africa (17) was adapted to this review. The An 80% cut-off was considered appropriate based on the fact that no subminimum criteria were applied due to the heterogeneous nature of LBP data and that the average methodological score of all studies was 83%.

### Analytical approach

From the Extracted data, the pooled point and twelve-month prevalence of LBP among working for Ethiopia population, as well as the 95% confidence interval (CI), were calculated for conducting meta-analysis. Heterogeneity between studies was assessed using the Cochrane Q and I^2^-statistics, which quantifies the proportion of variance explained by between-study heterogeneity. According to Higgins et al.[26] Heterogeneity was measured by I^2^ and divided into four categories, I^2^ <□25%, 25–49%, 50–75%, and□>□75% represents no, low, moderate, and high levels of heterogeneity, respectively. Sensitivity analysis was performed using a random-effect model. Funnel plot symmetry visualization followed by Begg’s rank correlation and Egger’s regression asymmetry test methods were performed to detect the existence of publication bias. The point and twelve-month prevalence estimate of each study with a 95% confidence interval were used to estimate pooled prevalence using the Der Simonian and Laird’s random-effects model. Furthermore, to assess the possible sources of variation, univariate meta-regression was conducted based on a year of publications. Data analysis was conducted using STATA software version 15.0. A significance level of 0.05 was considered for the P-value.

## Result

The comprehensive search for published epidemiological research in the musculoskeletal disorders in general regardless of etiology and LBP specifically conducted in Ethiopia regardless of period and yielded 719 hits. As the main objective of our current review focused solely on the prevalence LBP, studies which reported musculoskeletal disorders other than LBP and LBP related to underlying diseases (e.g. Rheumatology, Cancer…) were excluded. Studies that are not specifically conducted to report LBP but reported LBP prevalence were included. After removing duplicates and screening of titles and articles, 43 studies were selected for full-text review. Consequently, 13 eligible studies were included in this review (18-30). The literature search, selection, and reviewing process are depicted in the PRISMA flow diagram Figure 1.

**Figure 1:**
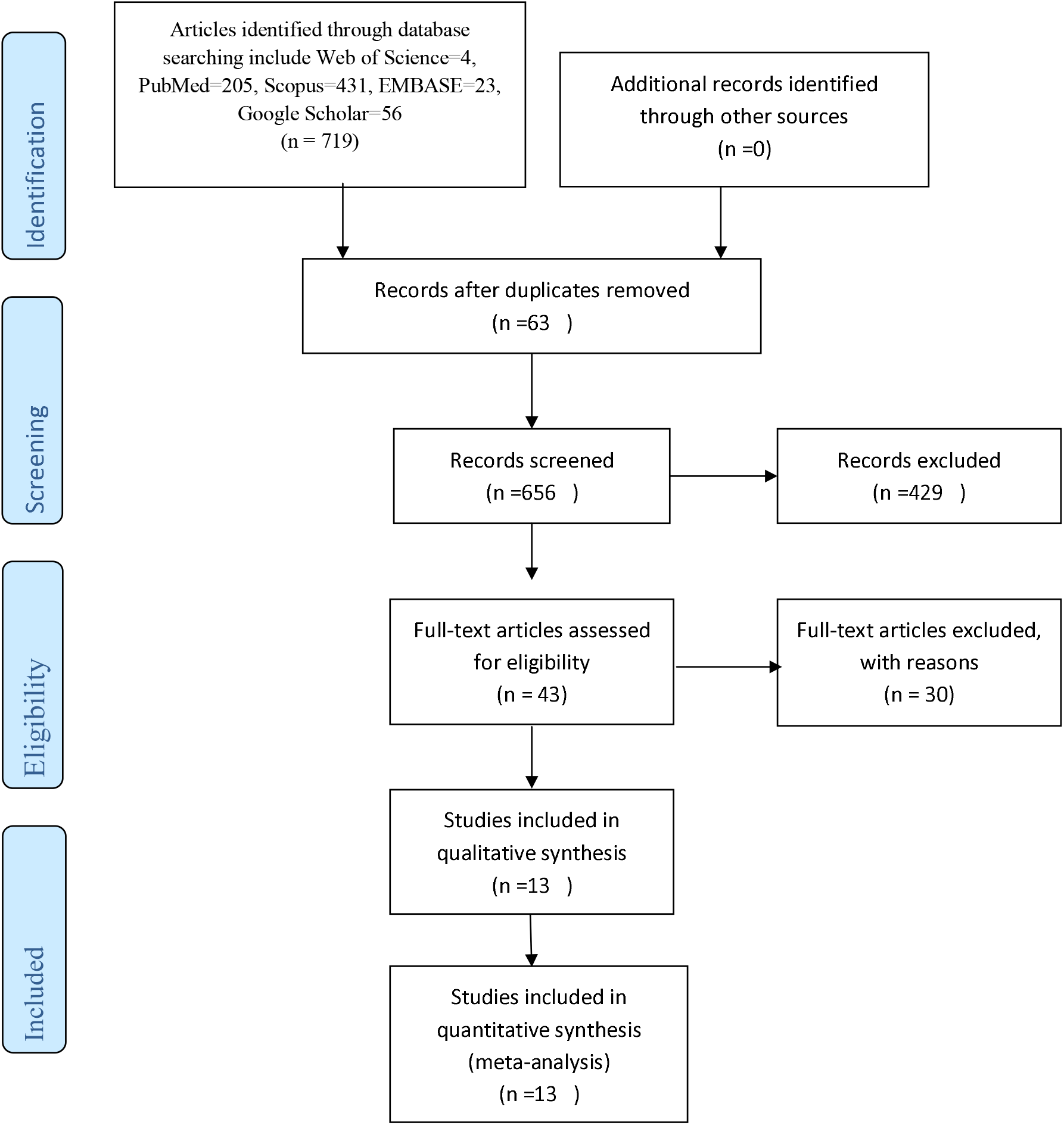
Flow chart of Studies selection

### General description of the studies reviewed

Descriptive data extracted from the thirteen included studies are reported as an overview summary (Table 1). The studies were published from 2013-2020. The sample size of the studies ranged from 300 to 824 individuals. Altogether, the thirteen studies included 6513 unique individuals, of whom 60% were males and 40% females. All studies reported response rate. The response rate varied from 90.4%-100% in the included studies. The mean response rates were 95.38%. Except for two studies (29, 30) which included only male subjects, the rest included both genders. The recall periods for LBP are reported as point and twelve-month prevalence. LBP prevalence is usually reported as the point or period prevalence. Point prevalence is measured at a single point in time (i.e., the number of people reporting LBP on the day of a survey) and Period prevalence is measured over a specified period, usually six months or 1 year (i.e., those people who report having had LBP in the past six-months or twelve-months) (31).

**Table 1:**
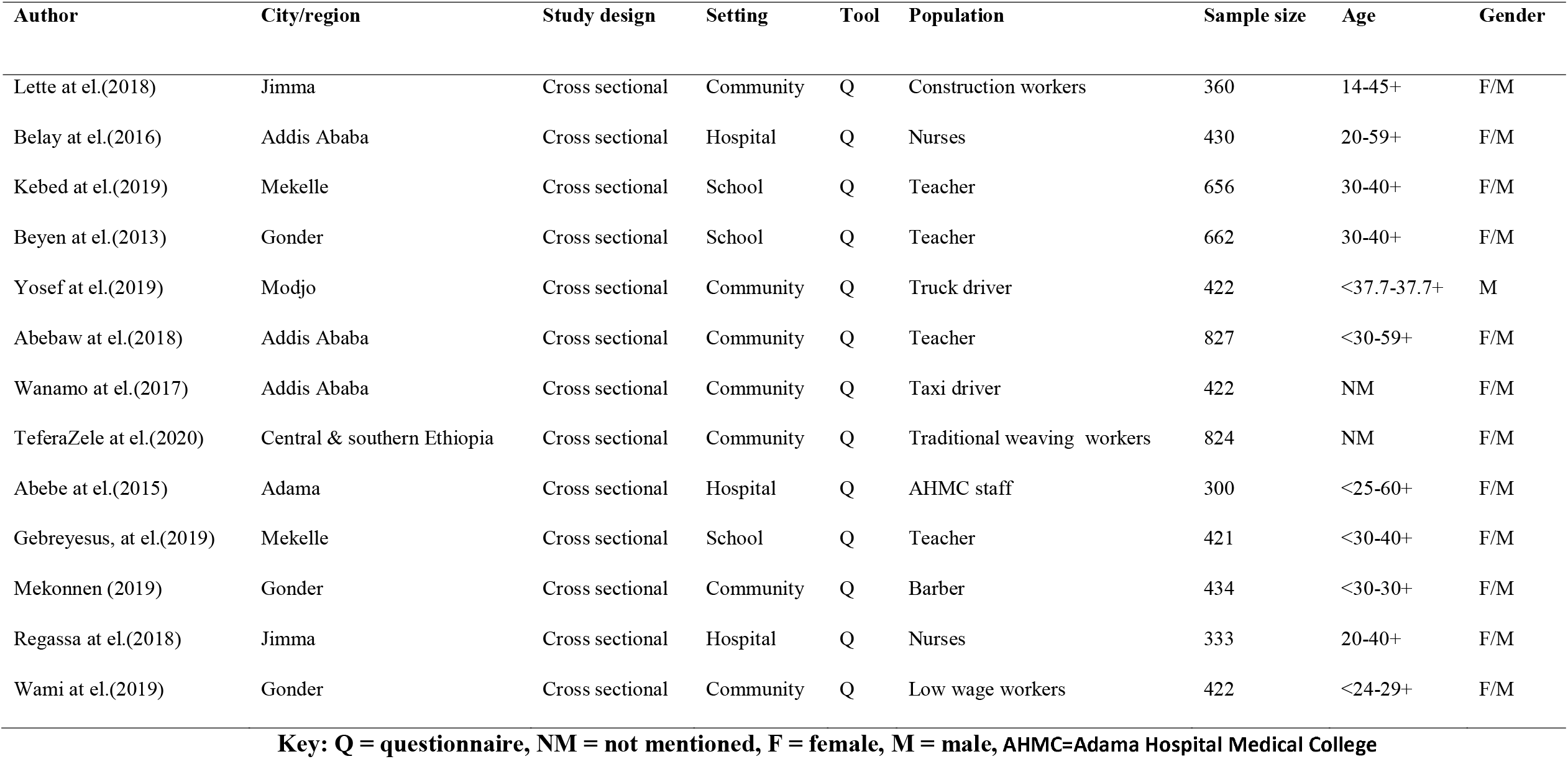
Study characteristics

### Methodological appraisal

The methodological quality scores of the included studies are reported in Table 2. All studies were descriptive with a quality score of higher than 80%. Ratings for each study were compared between the two evaluators, AGA and GA, with disagreement resolved by consensus.

**Table 2:** Methodological appraisal of included studies (n= 13)

| Author | 1 | 2 | 3 | 4 | 5 | 6 | 7 | 8 | 9 | 10 | Score (%) | MA |
| --- | --- | --- | --- | --- | --- | --- | --- | --- | --- | --- | --- | --- |
| Lette at el.(2018) | + | - | + | + | + | + | + | + | - | + | 80 | + |
| Belay at el.(2016) | + | - | + | + | + | + | + | + | + | + | 90 | + |
| Kebed at el.(2019) | + | - | + | + | + | + | + | + | + | + | 80 | + |
| Beyen at el.(2013) | + | - | + | + | + | + | + | + | - | + | 80 | + |
| Yosef at el.(2019) | + | - | + | + | + | + | + | + | + | + | 90 | + |
| Abebaw at el.(2018) | + | - | + | + | + | + | + | + | + | - | 80 | + |
| Wanamo at el.(2017) | + | - | + | + | + | + | + | + | + | + | 90 | + |
| TeferaZele at el.(2020) | + | - | + | + | + | + | + | + | - | + | 80 | + |
| Abebe at el.(2015) | + | - | + | + | + | + | + | + | - | + | 80 | + |
| Gebreyesus, at el.(2019) | + | - | + | + | + | + | + | + | + | + | 90 | + |
| Mekonnen (2019) | + | - | + | + | + | + | + | + | + | + | 90 | + |
| Regassa at el.(2018) | + | - | + | + | + | + | + | + | - | + | 80 | + |
| Wami at el.(2019) | + | - | + | + | + | + | + | + | - | + | 80 | + |

**Key: + = criteria fulfilled; – =criteria not fulfilled; MA=methodologically acceptable**

### Prevalence of LBP

Three studies reported on point prevalence of LBP (18, 20, 21). The point prevalence of LBP was estimated at 49% (95% CI 40; 58). Heterogeneity between the point prevalence of LBP was assessed and there was high heterogeneity (chi^230.5, d.f. =2, p=0.001, I^2 =93.44%). The summary analyses for point prevalence of LBP is depicted in Figure 2. Twelve studies reported on the twelve-month prevalence of LBP (19-30). The twelve-month prevalence of LBP was estimated at 56% (95% CI 49; 62). Heterogeneity between the twelve-month prevalence of LBP was assessed and there was high heterogeneity (chi^2= 298.06, d.f. =11, p=0.001, I^2 =96.3%). The summary analysis for the twelve-month prevalence of LBP is depicted in Figure 3.

**Figure 2:**
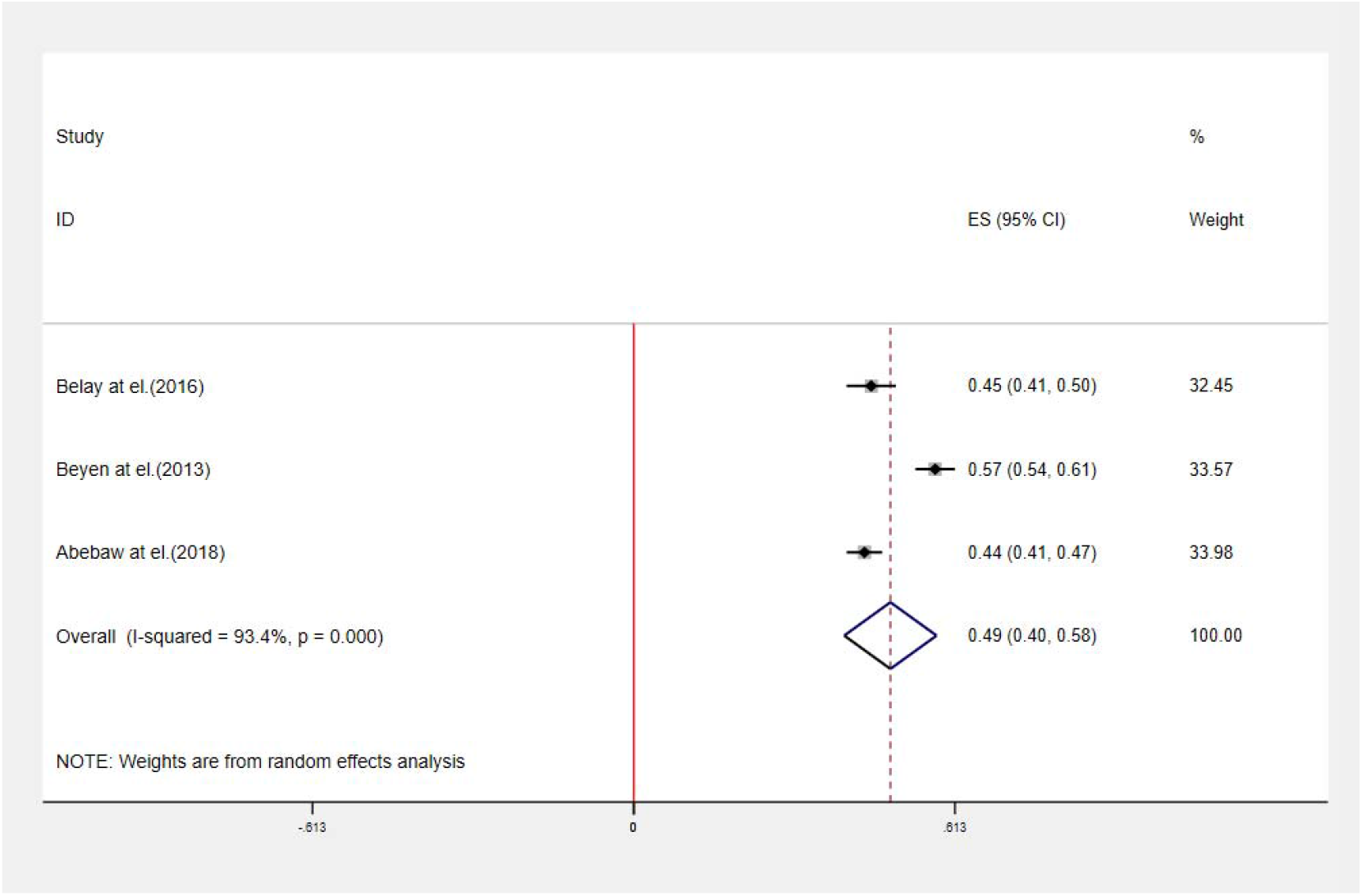
Pooled point prevalence of LBP (Random effects model used; ES- effect sizes, CI-Confidence interval).

**Figure 3:**
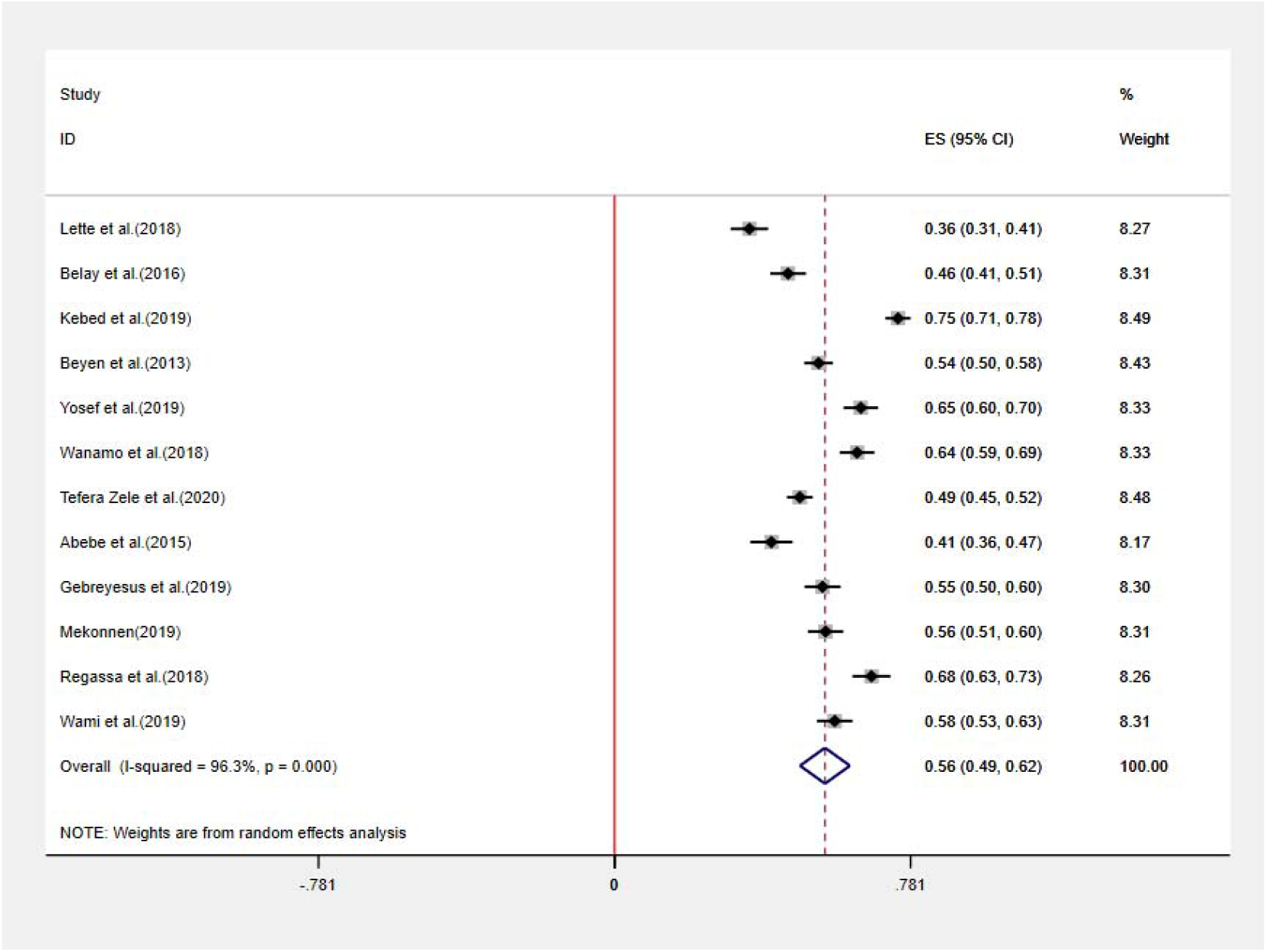
Pooled twelve-month prevalence of LBP (Random effects model used; ES- effect sizes, CI-Confidence interval)

### Socioeconomic and behavioral factors associated with low back pain

All studies included in this review (18-30) provided sufficient data to determine associations with potential risk factors. The main association factors were directly extracted from these publications and summarized in Table 3.

**Table 3:** A summary of socioeconomic and behavioral factors associated with low back pain among working Ethiopian population

| Population | Reference | Main Associated factors of low back pain |
| --- | --- | --- |
| Construction workers | (24) | Absence of vocational training, khat chewing, and working overtime |
| Nurses | (20) | Working shift, physical activities at work, sleep disturbance and felt little pleasure by doing things |
| Teachers | (23) | sleeping disturbance, prolonged standing, and irregular physical exercise |
| Teachers | (21) | Doing regular physical exercise, provisions of office at working institution and satisfaction with working environment and culture |
| Truck drivers | (30) | Smoking cigarette, physical inactivity, chronic diseases other than LBP, frequent lifting or carrying heavy objects, and perceived improper sitting posture while driving, perceived job |
| Teachers | (18) | Ergonomics training, enough teachers in school, prolonged sitting posture and poor work place social environment |
| Taxi drivers | (29) | Driving over ten years, uncomfortable seat, Frequency of lifting loads, Previous job that involved prolonged sitting, lack of physical exercise |
| Traditional weaving workers | (27) | Working seat without backrest , working 7 days in , workplace thermal level as warm, work at night and awareness on workplace safety as a protective effect |
| AHMC staffs | (19) | Obesity, stressed often, those worked in seated position more than six hours and those with long year experience |
| Teachers | (22) | Being female sex, smoking habit, doing regular physical and sleep disturbance |
| Barbers | (25) | Age, alcohol use, lack of safety training, working posture, and length of employment |
| Nurses | (26) | Lifting and transferring dependent patients, giving wound care, working in medical ward and Intensive care unit, working in mal-positions, working in the same positions for long period of time working with disoriented patients and bending or twisting back during work |
| Low wage workers | (28) | Being temporary employee, type of job which requires reaching/overstretching, engaging in a job that requires repetitive bending, making > 30 beds per day |

### Sensitivity analysis

Sensitivity analysis of LBP among the working Ethiopia population was performed using a random-effect model. Sensitivity analysis was carried out by excluding each study step-by-step from the meta-analysis and comparing point prevalence estimate before and after removing a single study. Accordingly, removing a single study did not alter the pooled prevalence estimate considerably, with sensitivity analysis ranging from 53.75% (when(23) was removed) and 57.34% (when(24) was removed) Figure 4.

**Figure 4:**
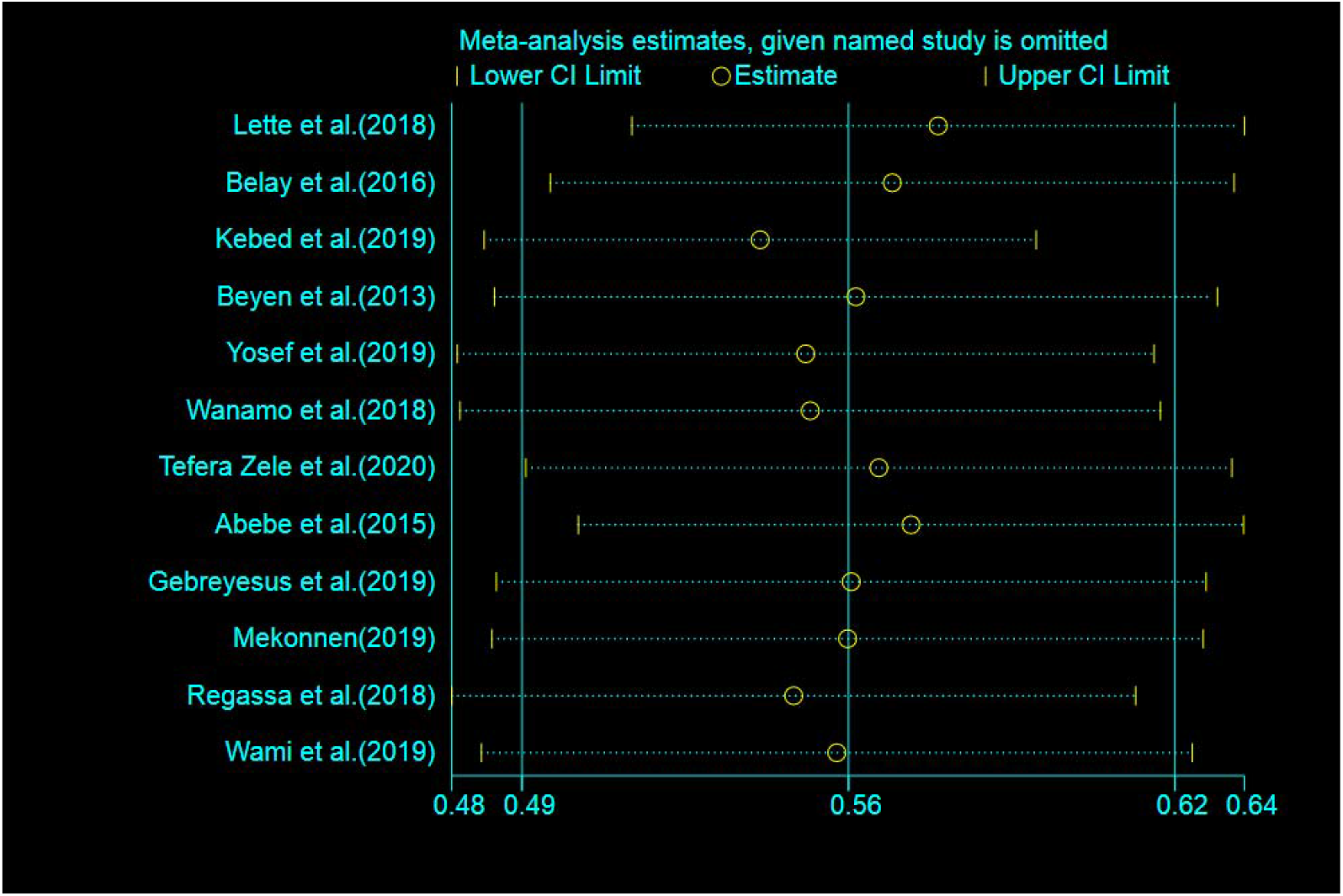
Sensitivity analysis of included studies to determine Prevalence of low back pain among working Ethiopian population.

### Meta-regression

Meta-regression analysis showed that there was no significant statistical relationship between the year of publication and the prevalence of the LBP (P-value = 0.345) Figure 5.

**Figure 5:**
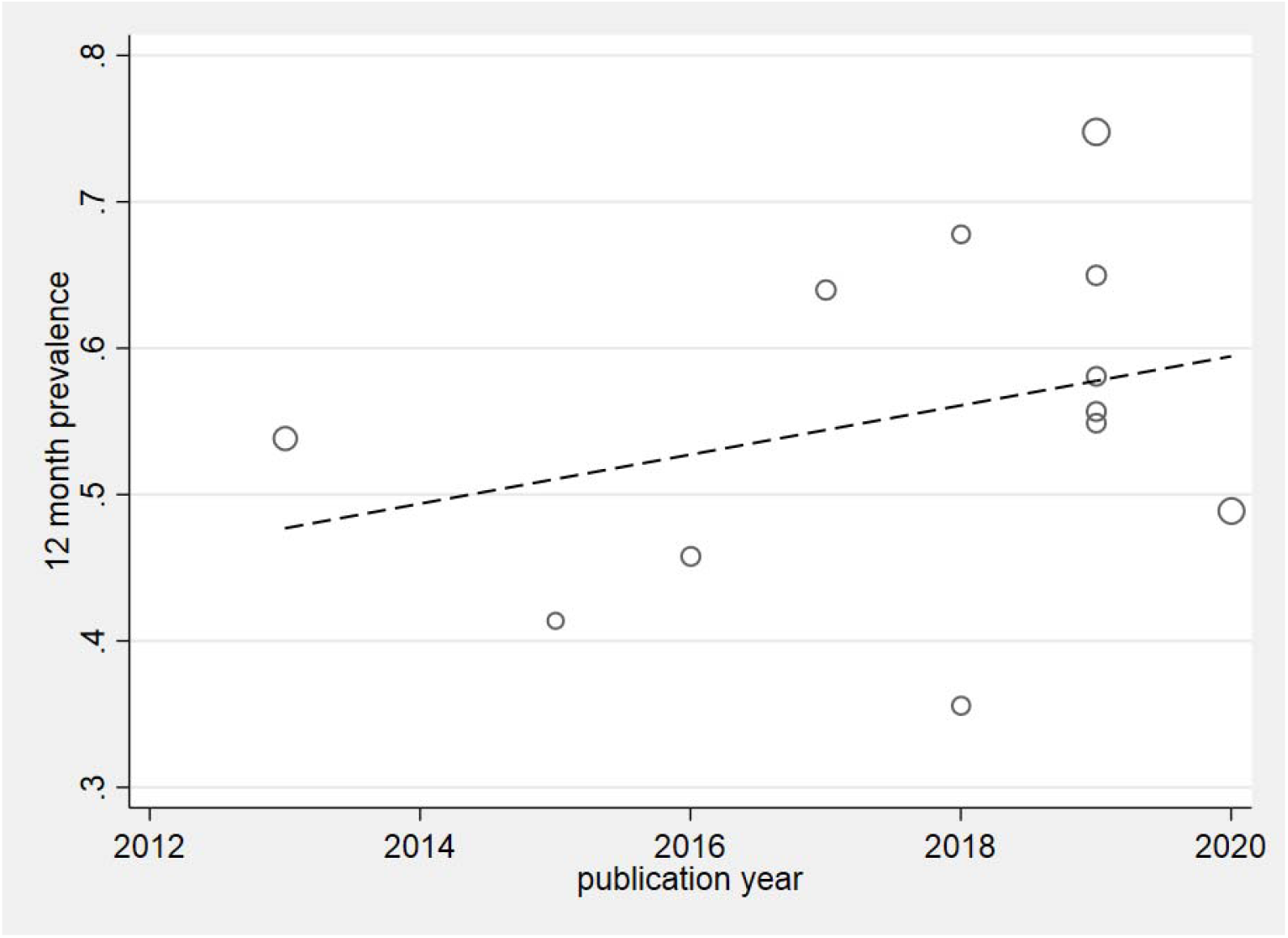
Meta-regression plot of Prevalence of low back pain among working Ethiopian population based on year of publication.

**Figure 6:**
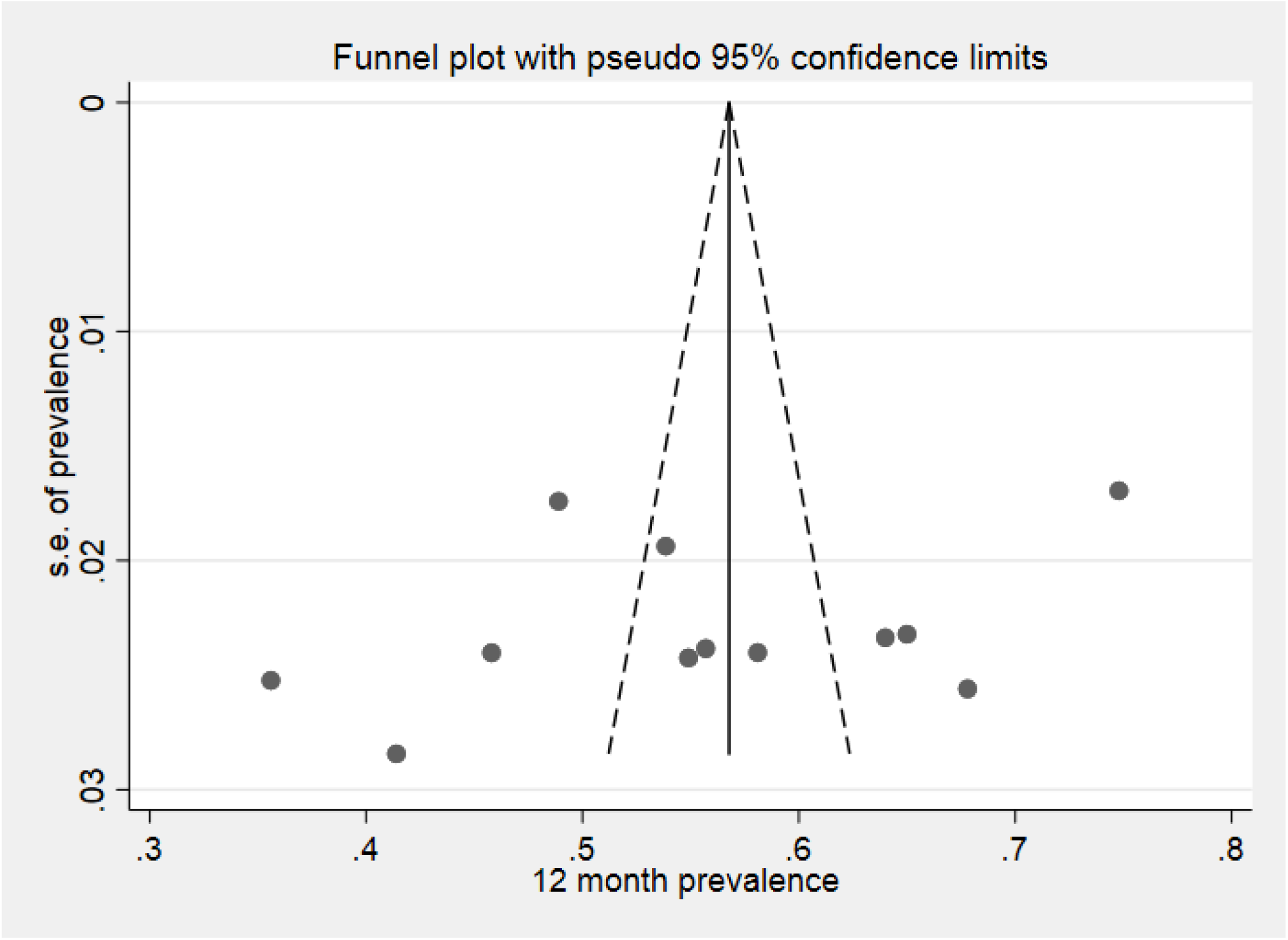
Meta funnel plot presentation of Prevalence of low back pain among working Ethiopian population. Abbreviation: s .e = standard error.

### Publication bias

The funnel plot symmetry visual inspection was used to assess the presence of publication bias qualitatively. This identified that the publication bias was not significant (figure.6). Besides, the absence of publication was statistically confirmed by Egger’s weighted regression test (bias coefficient (B)□=□-11.87, (95%CI□=□−□32.5–8.76%; p=□0.229) and Begg’s rank test (p-value of bias= 0.244). Therefore, we did not perform Trim and Fill analysis to adjust the final pooled prevalence estimate.

## Discussion

Meta-analyses of the observational data collected from the eligible studies provide a summary estimate of the point and twelve-month prevalence. Among those studies that reported point prevalence, LBP prevalence ranged from 44% to 58%. And among those provided twelve-month prevalence, LBP prevalence ranged from 36% to 75%.

Extracted data showed that Teachers had the highest rate in both point and twelve-month prevalence (21, 23), possibly due to prolong standing during sessions in addition to other potential risk factors. Point prevalence and twelve-month prevalence of LBP in working Ethiopia population was found to be higher than recently reported estimates of global LBL prevalence in the general population (12, 33, 34). A systematic review that included general population studies published in 2012 reported the global prevalence of LBP that was designed from a total of 165 studies conducted from 54 countries around the globe, published between 1980 and 2009(33). In our review, the point prevalence of LBP was estimated at 49% (95% CI 40; 58), which is considerably higher than both the Global LBP prevalence estimate (18.3%) reported by Hoy et al. (33) and the Prevalence of LBP among Africans estimate (39%) reported by Morris et al (17). Twelve-month prevalence of LBP was estimated at 56% (95% CI 49; 62), which is substantially higher than the Global LBP prevalence estimate (38.5%) reported by Hoy et al. (33) but relatively equal to the prevalence of LBP among Africans estimate (57%) reported by Morris et al (17).

The summary estimates from our current review were compared in particular to North American and Western European countries. It was found that the point LBP prevalence was considerably higher than estimates provided for Canada (28.7%), Denmark (12–13.7%) and Sweden (23.2%), and was comparable to Germany (39.2%) and Belgium (33%)(34). Twelve-month prevalence of LBP was considerably higher than Spain (20%), and on par with Denmark (56%) and Ukraine (50.3%) (34). The pooled prevalence LBP for both point and twelve-month prevalence were compared to particularly LBP prevalence among the working population reported in developed countries such as the United States of America (25.7%), Canada (28%) and United Kingdom (18%) (35), and our estimate found to be much higher than these countries. The findings of our review, therefore, reiterate the fact that LBP is a burden and is therefore a major public health concern among developing nations (12, 36, 37). Despite the high burden, LBP lacks enough attention amid infectious diseases and epidemics such as HIV/AIDS in developing countries (38). The successful development and implementation of strategies and policies to address the burden of LBP among the working population in Ethiopia or countries with emerging economies is therefore necessary (17, 39).

The review process highlighted a number of challenges related to conducting and pooling relevant epidemiologic data. Some of these study weaknesses are: First, significant heterogeneity has been noted among included studies. Thus, the pooled prevalence estimate should be interpreted cautiously. Second, only the English language was used to retrieve studies. Third, many of Ethiopian LBP studies are published in local journals or as a postgraduate thesis and not all Ethiopian universities may have information technology systems which allow online access to their postgraduate thesis, which leads to difficulty in publishing, as well as access and retrieving such publications. However, this study has several strengths that need to be mentioned: I) PRISMA guideline has been strictly followed, II) Large sample sizes have been included to estimate pooled prevalence, III) Publication bias assessment and sensitivity analysis have also been conducted to ensure the robustness of study, IV) Evaluation of possible source of heterogeneity and trend analysis was also done.

## Conclusion

The results showed the high prevalence of LBP among working Ethiopian population, especially among Teachers. We believe that Prevention strategies addressing the early onset of LBP among the working population would most likely be the answer to the burden of LBP on future economies in Ethiopia

## Data Availability

Data will be made available upon request/reasonable request.

## Abbreviations

LBP: Low back pain
GBD: Global Burden of Disease
DALYs: Disability adjusted life years
YLDs: Years lived with disability
HIV/AIDS: Human Immunodeficiency Virus/Acquired Immune Deficiency Syndrome
CI: Confidence interval

## Declarations

### Ethics approval and consent to participate

N/A

### Consent for publication

N/A

### Availability of data and materials

All data are included within the manuscript.

## Acknowledgements

Tehran University of medical sciences-International campus

